# Understanding the alcohol harm paradox: A multivariable Mendelian randomization approach

**DOI:** 10.1101/2024.07.30.24311209

**Authors:** Gemma Sawyer, Hannah Sallis, Marcus Munafò, Liam Mahedy, Jasmine Khouja

## Abstract

The alcohol harm paradox, whereby low socioeconomic position (SEP) groups experience greater alcohol-related harms at a given level of alcohol consumption, is not yet fully understood. In observational studies, key drivers are correlated and share similar confounding structures. We used multivariable Mendelian randomization (MVMR) to estimate the direct causal effect of alcohol (drinks per week) and education (years of schooling) on multiple health outcomes, accounting for the effect of the other. Previously published genome-wide association summary (GWAS) statistics for drinks per week and years of schooling were used, and outcome summary statistics were generated from individual-level data from UK Biobank (N=462,818). Inverse variance weighted analyses demonstrated evidence for direct effects of alcohol and education on liver diseases (alcoholic liver disease: alcohol OR = 50.19, 95% CI 19.35 to 130.21 and education OR = 0.27, 95% CI 0.14 to 0.53; other liver diseases: alcohol OR = 1.82, 95% CI 1.12 to 2.94 and education OR = 0.42, 95% CI 0.30 to 0.58), mental and behavioural disorders due to alcohol (alcohol OR = 12.89, 95% CI 7.46 to 22.27 and education OR = 0.51, 95% CI 0.35 to 0.75), and stroke (alcohol OR = 1.94, 95% CI 1.30 to 2.89 and education OR = 0.73, 95% CI 0.55 to 0.97). There was evidence for direct effects of education on depression, anxiety, influenza/pneumonia, and heart disease. In contrast, there was evidence of total (without considering the effect of education), but not direct, effects of alcohol on depression, influenza/pneumonia, epilepsy, and injuries. Although caution is required when interpreting these results, given weak instruments for alcohol, these results provide some evidence that the alcohol harm paradox is partially due to the protective effect of additional years of education. Replication with strong genetic instruments for drinks per week would be necessary to draw causal inferences.

**Author Summary:** Individuals from lower socioeconomic position backgrounds tend to disproportionately experience alcohol-related physical and mental ill-health, despite reporting lower overall alcohol consumption than those from higher socioeconomic backgrounds. This is known as the alcohol harm paradox. One key difficulty in understanding this paradox is the methodological difficulty of establishing the relative contribution of multiple complex social behaviours. In this study, we used genetic variants associated with alcohol consumption and years of education to explore their direct effects on multiple health outcomes. The findings indicate that greater alcohol consumption and fewer years of education may each, independently increase the likelihood of developing various health conditions, including liver diseases, mental and behavioural disorders due to alcohol, and stroke. This may suggest that the alcohol harm paradox is due to the protective effect of additional years of education amongst those from higher socioeconomic position backgrounds, reducing their likelihood of developing the health conditions. However, these findings are preliminary and limited by various methodological issues, suggesting these findings should be interpreted with caution. Replication and further studies are needed.

## Introduction

Alcohol-related harms are common and represent a large burden of morbidity and mortality worldwide, contributing to 5.3% of all deaths and 5.1% of global burden of disease [1]. Moreover, such alcohol-related harms tend to be disproportionately experienced by individuals from low socioeconomic position (SEP) backgrounds, despite reporting lower alcohol consumption compared to people from high SEP individuals background, a phenomenon known as the alcohol harm paradox [2–5]. Despite evidence for this paradox, there is limited understanding of its origins.

There have been multiple explanations proposed for the alcohol harm paradox. One is that, whilst low SEP individuals report consuming less alcohol on average, other patterns of drinking behaviour are more common. For example, low SEP individuals are more likely to be in the extreme ends of drinking consumption, with a greater proportion abstaining or drinking very lightly compared to high SEP groups, but also being more likely to drink excessively or binge drink [6]. The possibility that drinking patterns may account for the alcohol harm paradox has been supported by research demonstrating a greater socioeconomic gradient for acute health outcomes that are wholly attributable to alcohol, compared to chronic conditions that are influenced by a wide range of behaviours [7]. An alternative explanation is that individuals of low SEP may be more likely to engage in other unhealthy behaviours, such as tobacco use, which are associated with multiple adverse health outcomes [8]. Observational studies have supported this explanation, providing evidence that individuals of lower SEP are increasingly likely to smoke [3]. Furthermore, adjusting for tobacco smoking reduces the association between low SEP and hospital admissions [9], while causal mediation analysis demonstrated that 18% of the total effect of income on alcohol-related mortality was due to indirect effects via smoking and BMI [10]. Whilst these proposed explanations have received some support from observational studies, they are limited in their ability to draw causal inferences and are yet to fully account for the alcohol harm paradox.

The primary complication with disentangling the alcohol harm paradox is that a combination of highly correlated causes, including alcohol consumption, SEP, and smoking, could all be important independent drivers. Therefore, it is necessary to establish the relative causal contribution of alcohol consumption alongside these other factors on the likelihood of developing multiple adverse health outcomes with novel causal inference methods, such as Mendelian randomization (MR). MR relies on Mendel’s law of random assortment, using genetic variants associated with the exposure as instrumental variables to assess the total causal effect of the exposure on the outcome [11]. Multivariable MR (MVMR) builds upon this method, using genetic variants associated with multiple exposures to estimate the direct causal effect of each exposure on the outcome, accounting for the effect of the other exposures [12]. The validity of causal inferences drawn using MVMR relies on three key assumptions [13]: (1) each exposure is robustly predicted by the genetic instruments; (2) there are no confounders of the outcome and any of the genetic instruments; and (3) the genetic instruments only impact the outcome through at least one of the exposures.

The current study applies this MVMR framework to the alcohol harm paradox to examine the direct causal effect of alcohol consumption on adverse health outcomes whilst accounting for education (as a proxy for SEP), and vice versa. We first conducted MR to examine the total causal effect of alcohol and education separately on adverse health, followed by MVMR to examine their direct (i.e., unique) causal effects. Comparison of the MR and MVMR results will provide insights into the relative contribution of alcohol and education. For example, if there appears to be a causal effect of alcohol in both the MR and MVMR, this suggests that alcohol consumption impacts the health outcome even when the impact of education is accounted for. However, if a causal effect of alcohol in the MR is attenuated in the MVMR, this may indicate that the impact of education was driving the evidence of a causal effect of alcohol consumption in the MR and imply that an alternative explanation is needed to account for the associations reported between alcohol consumption and the health outcome.

## Materials and Methods

The analysis plan for this study was pre-registered on the Open Science Framework (https://doi.org/10.17605/OSF.IO/5G8W4), and the methods presented here are consistent with this unless otherwise specified. We have followed the STROBE-MR guidelines in the reporting of this study (Supplementary Materials page 3-6) [14,15].

### Exposures

Summary-level data reported by the two Genome-Wide Association Studies (GWAS) were utilised to identify suitable genetic instruments associated with the relevant exposure phenotypes: drinks per week and years of schooling. In contrast with our pre-registration, the main analysis reported here focuses solely on these two exposures and additional supplementary analysis included cigarettes per day as a third exposure (see Supplementary Methods for details). We focus on the two exposures of drinks per week and years of schooling because alcohol and smoking are highly correlated and both suffer from weak instrument bias in the three-exposure MVMR, making it difficult to draw conclusions from results using this approach. To aid interpretation, the analysis comprised of a two-exposure model to explore whether SEP plays a causal role in the alcohol harm paradox. Consequently, the results of this study should be considered preliminary.

#### GWAS & Sequencing Consortium of Alcohol and Nicotine Use (GSCAN)

Liu and colleagues [16] report summary-level statistics from a GWAS meta-analysis of alcohol use based on data from participants of European ancestry from 29 cohorts (N=941,280). Alcohol use was defined as the reported number of drinks (combined across all alcohol types) consumed per week amongst active drinkers. Where individual studies recorded categorical bins of drinks per week (e.g., 1-4, 5-9), the midpoint value was taken. To minimise the impact of outliers, this was subsequently left-anchored and log transformed and, as a result, we can only infer direction of effect and not size of the effect. Analyses adjusted for age, sex, age and sex interaction, and 10 genetic principal components. Independent variants were reported to be associated with drinks per week if they met the standard significance threshold (p < 5×10^8^). The associated SNPs explained 2.5% of the variance in drinks per week. We utilised the publicly available summary statistics from this GWAS, excluding UK Biobank, to minimise sample overlap, and 23andMe, due to data sharing restrictions (N=226,223). Smoking (cigarettes per day) was also measured in this GWAS, as described in the Supplementary Material (page 1-2).

#### Social Science Genetic Association Consortium (SSGAC)

Okbay and colleagues [17] report summary-level statistics from a GWAS meta-analysis of education, defined as years of schooling, based on data from participants of European ancestry from 65 cohorts (N=293,723). Reported number of years of schooling was utilised where this was available. Where individual studies reported qualifications, the International Standard Classification of Education (ISCED; 1997) [18] was used to map each major qualification that could be attained in each country into one of seven categories: pre-primary education, primary education, lower secondary education, upper secondary education, post-secondary non-tertiary education, first-stage tertiary education, and second-stage tertiary education. This was subsequently utilised to impute a years of schooling value for each category. Analyses adjusted for age, sex, age and sex interaction, ten genetic principal components, and cohort-specific controls. Variants were reported to be associated with years of schooling if they met the genome-wide significance threshold (p <5×10^8^) and were independent.

### Health outcomes

Individual-level data, including genetic and outcome data for all individual participants, was obtained from UKB to generate summary statistics for multiple health outcomes.

*UK Biobank.* UK Biobank is a population-based cohort, which recruited 500,000 people aged between 40-69 years in 2006-2010. With their consent, they provided detailed information about their lifestyle, physical measures and had blood, urine and saliva samples collected and stored for future analysis. Using this individual-level data, we generated summary statistics by regressing each available SNP on each health outcome through the MRC-IEU UK Biobank GWAS pipeline [19] using BOLT-LMM (a linear mixed model) to account for relatedness and population stratification. Furthermore, we additionally adjusted for age, sex, and genotyping chip array. For the main two-exposure MVMR analysis, this was conducted for the overall UKB sample; however, it was also conducted in four smaller samples stratified by smoking status for comparison with the supplementary three-exposure MVMR: ever smokers (defined as those reporting smoking 2 or more cigarettes), never smokers (defined as those reporting smoking 1 or less cigarettes), current smokers, and former smokers (Figure 1).

**Figure 1.**
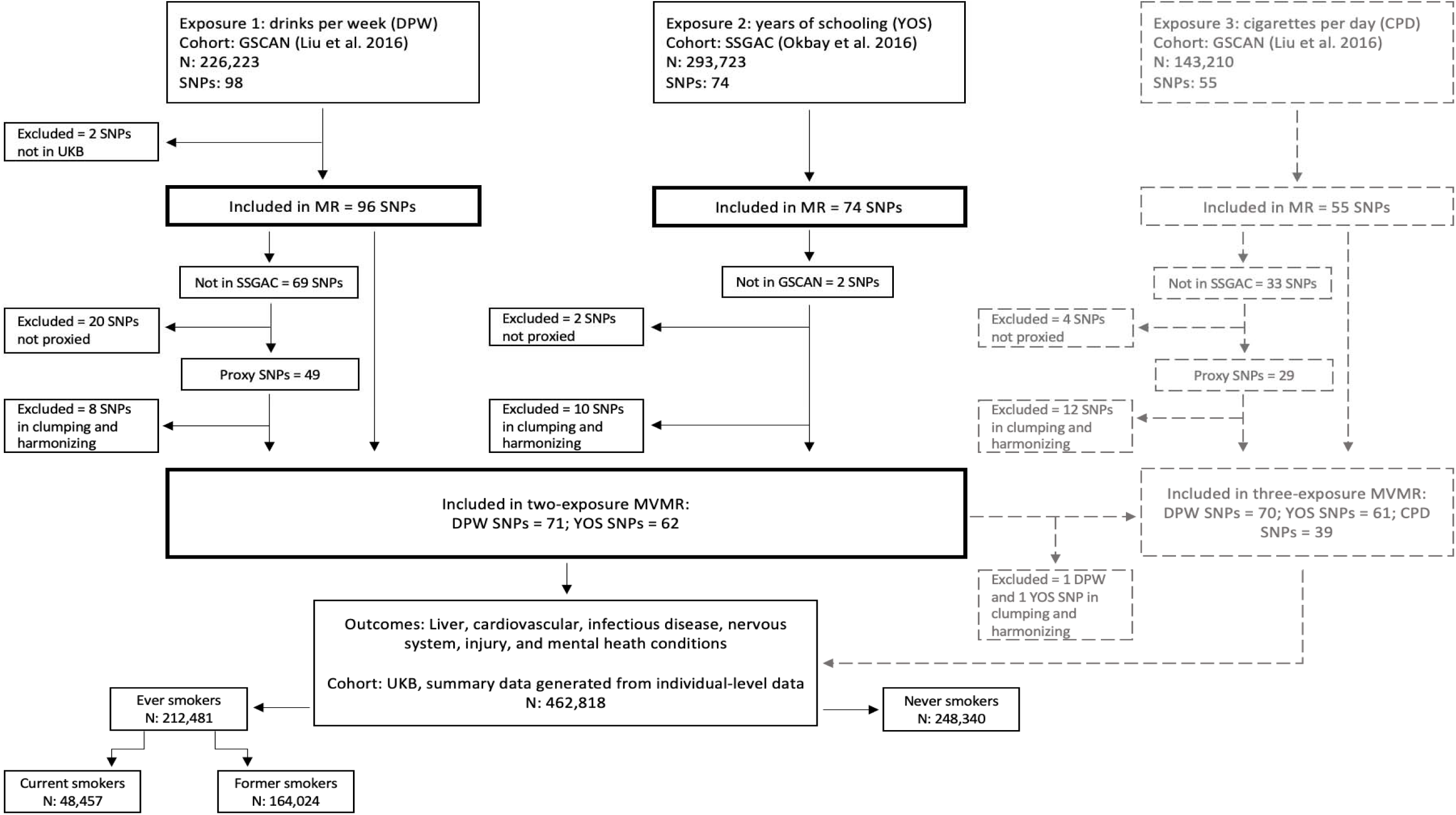
Flowchart demonstrating the source of exposure and outcome data and the inclusion and exclusion of genetic variants (single nucleotide polymorphisms; SNPs) in this analysis. Details of the three-exposure MVMR (grey dashed lines) are in the Supplementary Materials.

Our pre-registered analysis specified 15 outcomes wholly or partially attributable to alcohol, based upon ICD-10 summary diagnoses classification codes (UKB field ID 41270; see Supplementary Table S1). However, once all the necessary exclusions had been made, four outcomes (alcoholic cardiomyopathy, alcoholic myopathy, alcoholic polyneuropathy, and alcoholic poisoning) had a case number below 50 or less than 0.1% in at least one of the stratified datasets and were therefore removed from the analysis. The remaining outcomes included alcoholic liver disease, other liver diseases, stroke, ischaemic heart disease (IHD), influenza/pneumonia, viral hepatitis, epilepsy, injuries, mental and behavioural disorders due to alcohol, depression, and anxiety.

### Analysis

Analyses were conducted in R version 4.1.2 [20], using the *Mendelian Randomization* [21], *TwoSampleMR* [22]*, MVMR* [23] and *MR-PRESSO* [24] R packages.

#### Selection of genetic variants

Single nucleotide polymorphisms (SNPs) were selected for inclusion in the analysis if they were reported to be associated with the exposure phenotype of interest (alcohol use or education) and (conditionally) independent of any other SNP associations at the genome-wide significance level (p<5×10) in the relevant GWAS. In GSCAN, this involved sequential forward selection whereby SNPs within each locus were iteratively included in the set of independent SNPs until a SNP was no longer significant using a partial correlation-based score statistic whereas SSGAC applied conditional joint analysis in the Genome-Wide Complex Trait Analysis software (GCTA-COJO) which selected SNPs through a stepwise conditional regression model. These SNPs also needed to be available in UK Biobank, resulting in 96 drinks per week and 74 years of schooling SNPs in the univariable MR. For the MVMR analyses, where a SNP from GSCAN (drinks per week) was not available in SSGAC (years of schooling), we selected proxy SNPs with a minimum linkage disequilibrium (LD) R of 0.8. Moreover, an additional clumping stage was conducted in the MVMR to ensure all exposure SNPs were independent (LD R^2^ < 0.1, clumping window > 500 kb), resulting in 71 drinks per week and 62 years of schooling SNPs in the MVMR (Figure 1; see Supplementary Table S2 for details).

We assessed instrument strength using a conditional F-statistic and instrument validity using Cochran’s Q-statistic. In line with conventional guidelines, the F-statistic should be greater than 10 and the Q-statistic should be less than the number of SNPs [25].

#### Univariable MR

We estimated the total causal effects of alcohol use and education on all 11 health outcomes using two-sample, summary level MR. The main analysis was conducted using inverse variance weighted (IVW) regression [26]. We also conducted sensitivity analyses using three complementary MR methods MR-Egger [27], weighted median estimation [28], and weighted mode estimation [29]. These methods hold different assumptions regarding pleiotropy and therefore consistent results across the methods provide stronger evidence that the observed effects are causal.

#### Multivariable MR

In MVMR analyses, we estimated the direct causal effect of alcohol use, education, and tobacco use on all health outcomes using two sample, summary level MVMR with two complimentary methods – MVMR-IVW and MVMR-Egger. Whilst we intended to conduct MVMR-PRESSO in our pre-registered analysis plan, we had to remove this method due to issues with the R package.

## Results

The MR-IVW, MVMR-IVW, MR-Egger and MVMR-Egger results exploring the total and direct effects of alcohol and education are displayed in Figure 2. The complete results of the MVMR analyses exploring the direct effects of (1) alcohol and education (n = 133 SNPs) and (2) alcohol, education, and smoking (n = 171 SNPs) on health outcomes are displayed in Supplementary Tables S3-S13. The results are presented in odds ratios (ORs) per standard deviation (SD) increase in either drinks per week or years of schooling.

**Figure 2.**
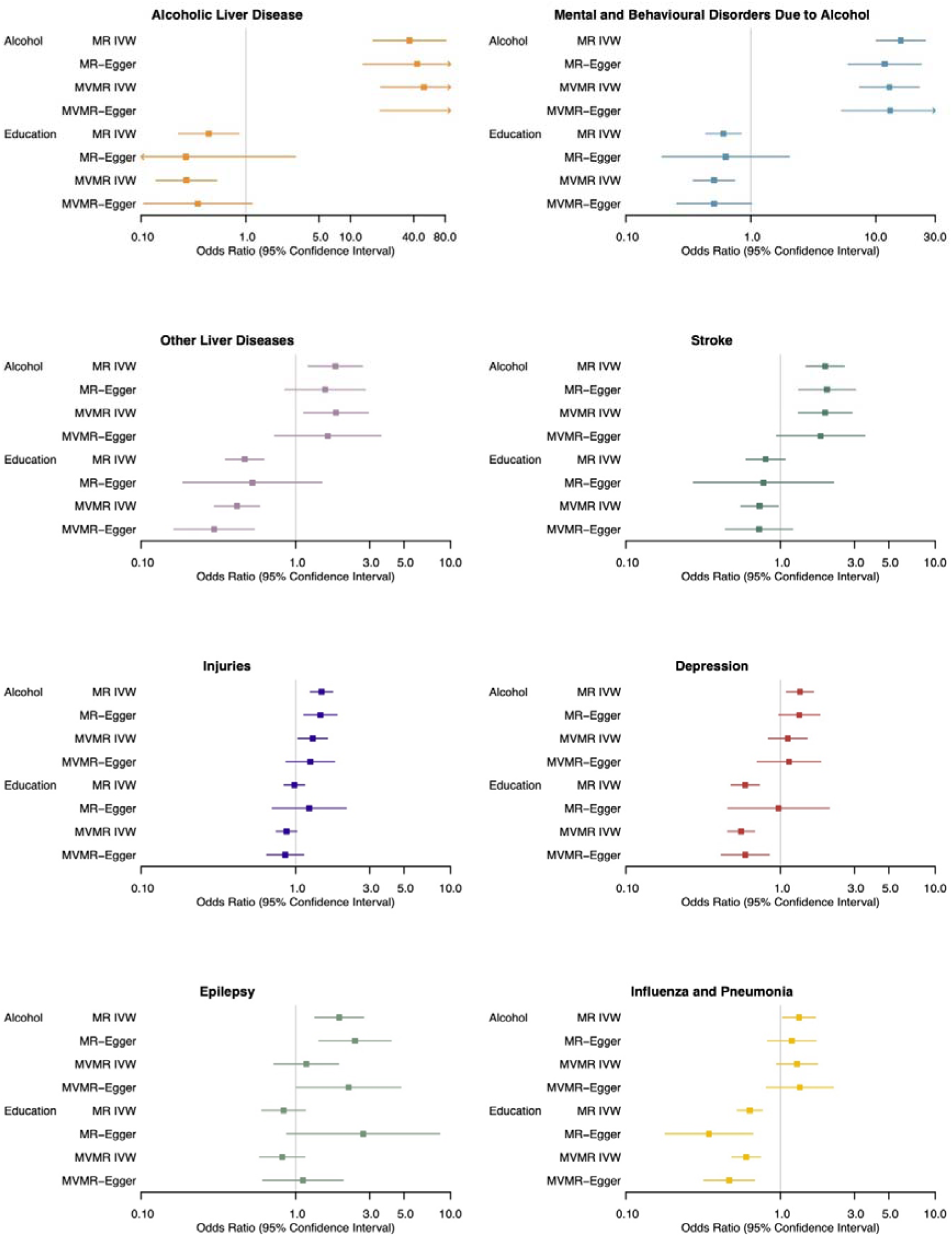
Forest plots displaying the effect of alcohol (drinks per week) and education (years of schooling) on eight health outcomes using Mendelian randomization (MR) and multivariable MR (MVMR) analysis: inverse variance weighted (IVW) and Egger regression.

### Instrument strength and heterogeneity

Instrument strength was calculated using the conditional F-statistic. In the MR, the F-statistics suggest that the SNPs included in the analysis are strong instruments for alcohol (F = 14.87) and education (F = 35.32). The conditional F-statistics in the two exposure MVMR indicate that the included SNPs are strongly associated with education whilst accounting for the effects of alcohol (F = 17.42), but not strongly associated with alcohol while accounting for education (F = 6.32).

The Cochran’s Q statistics were greater than the number of SNPs included in most of the models, indicating heterogeneity. Therefore, the results also include MVMR-Egger, which provides estimates that are robust to directional pleiotropy under the assumption that pleiotropy is uncorrelated with the strength of the association between the SNP and exposure. However, MVMR-Egger analyses have reduced statistical power compared with IVW analyses and therefore we must consider the results from both analysis when drawing conclusions. Where Egger results indicate a similar effect size but with larger confidence intervals which cross the null, we interpret this as evidence in support of the IVW analysis.

### Alcoholic liver disease

There was evidence for a total (OR = 36.57, 95% CI 16.46 to 81.24) and direct (OR = 50.19, 95% CI 19.35 to 130.21) effect of alcohol on alcoholic liver disease when accounting for education. There was also evidence that education decreased the odds of alcoholic liver diseases in the univariable (OR = 0.44, 95% CI 0.23 to 0.86) and multivariable (OR = 0.27, 95% CI 0.14 to 0.53) analysis when accounting for alcohol (Figure 2; Supplementary Table S3). Whilst the MR-Egger and MVMR-Egger results were consistent with the IVW results for alcohol, for education, the effect estimates were similar but the wider confidence intervals overlapped the null.

### Mental and behavioural disorders due to alcohol

There was evidence for a total (OR = 15.88, 95% CI 10.08 to 25.01) and direct (OR = 12.89, 95% CI 7.46 to 22.27) effect of alcohol on mental and behavioural disorders due to alcohol, as well as a total (OR = 0.60, 95% CI 0.44 to 0.83) and direct (OR = 0.51, 95% CI 0.35 to 0.75) effect of education on mental and behavioural disorders due to alcohol (Figure 2; Supplementary Table S4). The MR- and MVMR-Egger findings were consistent with the IVW findings for alcohol and showed similar effect estimates for education, although the confidence intervals crossed the null.

### Other liver diseases

There was evidence for a total (OR = 1.80, 95% CI 1.21 to 2.70) and direct (OR = 1.82, 95% CI 1.12 to 2.94) effect of alcohol on other liver diseases, as well as evidence for a total (OR = 0.47, 95% CI 0.35 to 0.62) and direct (OR = 0.42, 95% CI 0.30 to 0.58) effect of education on other liver diseases (Figure 2; Supplementary Table S5). The MR- and MVMR-Egger results calculated similar effect estimates, although the wider confidence intervals crossed the null for alcohol and the total effect of education.

### Stroke

There was evidence for a total (OR = 1.94, 95% CI 1.46 to 2.58) and direct (OR = 1.94, 95% CI 1.30 to 2.89) effect of alcohol on stroke, when accounting for education. Also, whilst there was no clear evidence for a total effect of education on stroke, there was some evidence of a direct effect in the MVMR when accounting for alcohol (OR = 0.73, 95% CI 0.55 to 0.97) (Figure 2; Supplementary Table S6). The MR- and MVMR-Egger findings were similar to the IVW findings in terms of the effect estimates. However, the confidence intervals were wider and therefore crossed the null for education and the direct effect of alcohol.

### Injuries

There was evidence for a total (OR = 1.47, 95% CI 1.24 to 1.73) and direct (1.29, 95% CI 1.03 to 1.60) effect of alcohol on injuries. However, there was no clear evidence for a total (OR = 0.98, 95% CI 0.85 to 1.14) or direct (OR = 0.87, 95% CI 0.75 to 1.02) effect of education (Figure 2; Supplementary Table S7). The MR- and MVMR-Egger results supported the IVW results for education and the total effect of alcohol; however, the confidence intervals crossed the null for the direct effect of alcohol.

### Depression

In the univariable MR, there was evidence for a total effect of alcohol on depression (OR = 1.33, 95% CI 1.09 to 1.64); however, there was no clear evidence for a direct effect when accounting for education (OR = 1.11, 95% CI 0.84 to 1.48). There was evidence that education decreased the odds of depression in the univariable (OR = 0.59, 95% CI 0.48 to 0.73) and multivariable analyses when accounting for alcohol (OR = 0.56, 95% CI 0.45 to 0.68) (Figure 2; Supplementary Table S8). The MR- and MVMR-Egger results supported the IVW findings of a total, but not direct, effect of alcohol (although the wider confidence intervals crossed the null), as well as a direct effect of education. However, the MR-Egger did not support IVW evidence for a total effect of education.

### Epilepsy

Whilst there was evidence for a total effect of alcohol on epilepsy (OR = 1.91, 95% CI 1.33 to 2.75), there was no clear evidence for a direct effect when accounting for education (OR = 1.17, 95% CI 0.72 to 1.89). Moreover, there was no clear evidence for a total (OR = 0.83, 95% CI 0.60 to 1.15) or direct (OR = 0.82, 95% CI 0.58 to 1.14) effect of education on epilepsy (Figure 2; Supplementary Table S9). The MR-Egger results supported a total effect of alcohol and also found some evidence for a direct effect, which was not indicated in the IVW findings. The MR- and MVMR-Egger effect estimates for education were in the opposite direction to the IVW results, although all confidence intervals crossed the null.

### Influenza and pneumonia

There was evidence for a total (OR = 1.32, 95% CI 1.03 to 1.68) but not a direct effect of alcohol on influenza/pneumonia when accounting for education (OR = 1.28, 95% CI 0.94 to 1.73). There was evidence that education decreased the odds of influenza/pneumonia in the univariable (OR = 0.63, 95% CI 0.53 to 0.76) and multivariable (OR = 0.60, 95% CI 0.48 to 0.74) analysis (Figure 2; Supplementary Table S10). The MR- and MVMR-Egger results were primarily consistent with the IVW results, supporting a total and direct effect of education and no direct effect of alcohol. Whilst the effect estimate for the total effect of alcohol was similar, the MR-Egger confidence interval crossed the null.

### Other outcomes

There was no clear evidence for a total or direct effect of alcohol on IHD (Supplementary Table S11), viral hepatitis (Supplementary Table S12), or anxiety (Supplementary Table S13). There was evidence for a total (OR = 0.61, 95% CI 0.53 to 0.70) and direct (OR = 0.62, 95% CI 0.52 to 0.74) effect of education on IHD when accounting for alcohol. There was also evidence for a total (OR = 0.67, 95% CI 0.52 to 0.86) and direct (OR = 0.62, 95% CI 0.48 to 0.80) effect of education on anxiety. There was no clear evidence for a total or direct effect of education on viral hepatitis.

Overall, the MR- and MVMR-Egger results were mostly similar to the IVW results for IHD, anxiety, and viral hepatitis. The only differences were that the MR-Egger did not support a total effect of education on IHD or anxiety.

## Discussion

We find evidence for direct effects of alcohol and education on alcoholic and other liver diseases, mental and behavioural disorders due to alcohol, and stroke, indicating that alcohol use increases the likelihood of such outcomes, whereas years of education decreases their likelihood. There was also evidence of a direct effect of education on depression, anxiety, influenza/pneumonia, and IHD. In contrast, there was evidence of a total effect, but not a direct effect (i.e., when accounting for education), of alcohol on depression, influenza/pneumonia, epilepsy, and injuries.

The direct effects of alcohol on liver diseases and mental and behavioural disorders due to alcohol are consistent with the aetiology of such conditions, as they are known to be ‘wholly attributable’ to alcohol [7,30]. For such ‘wholly attributable’ outcomes, these results are consistent with previous findings from observational studies suggesting the paradox may be due to the multiplicative contribution of drinks per week and years of schooling, which both may impact the likelihood of developing such health conditions [3,4]. Moreover, the direct effect of alcohol on stroke is supported by a previous MR meta-analysis demonstrating robust causal relationships between alcohol consumption and cardiovascular diseases, including ischaemic stroke [31].

However, the results for other outcomes (‘partially attributable’) imply that years of schooling, but not drinks per week, contribute to their development, including total but not direct effects of alcohol on some conditions such as depression, epilepsy, and injuries when accounting for education. This finding resonates with a recent genome wide meta-analysis which found novel genetic loci associated with problematic alcohol use and its comorbidities, such as depression and other mental health disorders [32], suggesting that the contribution of alcohol to these may be, at least partially, mediated by other correlated factors.

The evidence for a causal contribution of years of schooling is consistent with previous natural and genetic studies suggesting that raising the school leaving age and additional years of education reduce the likelihood of developing adverse health outcomes [33,34]. The protective effects of education observed in conditions such as depression, anxiety, and ischemic heart disease (IHD) are consistent with previous population-based observational studies highlighting the protective effect of education on anxiety and depression [35] and large-scale MR studies demonstrating that education positively impacts coronary heart disease [36].

The finding that drinks per week is not causally related to many of the ‘partially attributable’ conditions may be because drinks per week is a simplified a measure of alcohol consumption. Other, more complex drinking patterns (such as alcohol frequency, drinks consumed per drinking occasion, and alcohol type) may be more important in the development of these conditions. Previous observational research has reported that low SEP groups are more likely to binge drink or exceed high weekly drinking thresholds [6]. This has also been supported by genetic research which have reported opposite genetic correlations between SEP and drinking frequency compared with drinking consumption [37], and have demonstrated relationships between more years of education and reduced binge drinking, number of drinks per drinking occasion, and spirit intake using MR [38]. Therefore, the results for these ‘partially attributable’ conditions may be because their relationship with alcohol consumption is more nuanced than is being captured by the genetic instruments associated with drinks per week.

Overall, the present study adds nuance to the alcohol harm paradox by illustrating that education not only mitigates the adverse effects of alcohol but also plays an independent protective role in mental and physical health outcomes. This complements earlier genetic studies on both alcohol use and educational attainment, which emphasise the complex interplay between these factors and various health outcomes. However, it is important to be cautious and consider the strength of instruments when interpreting these results. The conditional F-statistics indicate that the instruments were not strongly associated with alcohol in the MVMR, despite strong associations in the MR. Therefore, we cannot be as confident or draw causal inferences from our estimates of direct effects; instead, our findings may be indicative of possible causal effects which would need to be replicated with strong instruments to be confident. In addition, we were not able to explore the relative contribution of tobacco use alongside alcohol consumption and SEP as initially intended due to the weak genetic instruments for CPD and, therefore, we can only consider these results as exploratory as they were not all specified in our pre-registered analysis plan.

### Strengths and Limitations

This is the first study, to our knowledge, to apply MVMR, a novel causal inference method, to understand the direct causal effects of alcohol and education on multiple alcohol-related conditions to enhance understanding of the alcohol harm paradox. Also, the large sample size of the GWAS for education will have enhanced statistical power and the conditional F-statistics suggest good instrument strength for years of schooling. However, this study also has multiple limitations.

As previously mentioned, the conditional F-statistics indicate that alcohol suffers from weak instruments in the MVMR, which may possibly bias our results. Heterogeneity is indicated by the Cochran’s Q statistics and, even though we applied MR-Egger to address this, heterogeneity may remain, indicating the analyses may also be biased by invalid instruments. Directional pleiotropy is similarly likely to persist in our analyses despite applying pleiotropy-robust methods as indicated by the MR-Egger intercepts, which are not null in most of the models. This suggests that the genetic instruments may influence the adverse health outcomes within this study via pathways that do not include alcohol or education and therefore may not tell us about the causal effect of alcohol or education on such outcomes. Moreover, drinks per week is a somewhat limited phenotype to reflect alcohol as many factors influence the effects of alcohol beyond number of drinks, including binge patterns and alcohol types. Similarly, education is one factor that contributes to an individual’s SEP; factors such as income, neighbourhood, and occupation are also important and may show different relationships. Therefore, focusing purely on drinks per week and years of schooling may be restricting our understanding. Combined with the exploratory nature of these analyses, these limitations suggest that caution is highly necessary in interpreting the current findings.

### Future Research and Implications

Due to limitations within the study regarding weak instruments and possible pleiotropy, the current study should be replicated with newly available GWAS data [39] allowing valid and stronger instruments to be utilised in these analyses. Replication will help confirm whether the results presented in this study reflect the true causal effects of alcohol and education or are due to methodological limitations. If the current findings were supported, this would suggest that the alcohol harm paradox may be due to the protective effect of years of schooling on many alcohol-related conditions resulting in a reduced likelihood of higher SEP groups developing such outcomes. Future research may therefore benefit from exploring the mechanisms through which education impacts such health outcomes to inform preventative policy. Beyond this, applying this MVMR framework to explore the contribution of alternative exposures (such as different drinking patterns, income, and impulsivity/risk-taking behaviour) may provide a useful next step for future research aiming to disentangle the alcohol harm paradox.

## Conclusion

This research demonstrates the utility of applying the MVMR framework to enhance understanding of complex behavioural phenomena. We found evidence for a direct effect of alcohol on alcoholic and other liver diseases, mental and behavioural disorders due to alcohol, and stroke. There was also evidence for a total effect of alcohol on depression, epilepsy, influenza/pneumonia, and injuries, which were attenuated when education was accounted for. Despite weak instrument bias, such results cautiously indicate that many partially alcohol-attributable conditions are causally impacted by years of education, which we use here as a proxy for SEP. Replication with strong instruments is necessary to confirm these findings. Future research may endeavour to explore the causal effect of education on other partially alcohol-attributable conditions and identify the mechanisms through which further years of education protects against these conditions.

## Data Availability

UK Biobank data is available at www.ukbiobank.ac.uk. GSCAN data is available at doi:10.1038/s41588-018-0307-5. SSGAC data is available at doi:10.1038/nature17671.

doi:10.1038/s41588-018-0307-5

doi:10.1038/nature17671

## Acknowledgements

This research has been conducted using the UK Biobank Resource under application number 9142. This work uses data provided by patients and collected by the NHS as part of their care and support.

