## Supplemental Material for "Understanding the alcohol harm paradox: A multivariable Mendelian randomization approach"

**Supplementary Material**

**Three-exposure MVMR including cigarettes per day.**

*Exposures*

Consistent with the pre-registered analysis plan, we conducted analyses with three exposures: drinks per week (DPW), years of schooling (YOS), and cigarettes per day (CPD).

We utilised publicly available summary statistics from the GWAS & Sequencing Consortium of Alcohol and Nicotine Use meta-analysis (GSCAN; Liu et al. 2019) based on data from participants of European ancestry from 29 cohorts (N=337,334). Current or former smokers reported their average number of cigarettes smoked per day. Where studies reported cigarettes per day as an integer, responses were categorised into one of five bins (1-5, 6-15, 16-25, 26-35, and 35+), whereas where studies had reported pre-defined bins, these were used. Analyses adjusted for age, sex, age x sex interaction, and ten genetic principal components and variants were reported to be associated with DPW if they met the standard significance threshold (5 x 10^8^). The associated SNPs explained 4% of the variance in CPD. We utilised the publicly available summary statistics from this GWAS, excluding UK Biobank (UKB) to minimise sample overlap and 23andMe due to data sharing restrictions (N=143,210). The summary statistics used to identify genetic instruments for DPW and YOS are described in the main text.

*Health outcomes*

The health outcomes were obtained from individual-level data from UKB as with the main analysis. We generated summary statistics by regressing each available SNP on each health outcome through the MRC-IEU UKB GWAS pipeline using a BOLT-LMM (linear mixed model) to account for relatedness and population stratification, adjusting for age, sex, and genotyping chip array. This was conducted in the entire sample, as well as in four smaller samples stratified by smoking status: ever smokers (defined as those reporting smoking 2 or more cigarettes), never smokers (defined as those reporting smoking 1 or less cigarettes), current smokers, and former smokers (see Figure 1 in main text). The health outcomes were the same as in the main analysis.

*Analysis*

The analysis procedure was identical to the main analysis. SNPs were selected for inclusion if they were associated with the exposure phenotype of interest, conditionally independent of any other SNP associations at the genome-wide significance level (p < 5×10^-8^), and available in UKB. This resulted in 55 CPD SNPs in the univariable MR. The DPW and YOS univariable MR analyses are outlined in the main text. For the MVMR, proxy SNPs were identified with a minimum linkage disequilibrium (LD) R^2^ of 0.8 when a CPD SNP was not available for YOS. An additional clumping stage was also conducted to ensure all exposure SNPs were independent (LD R^2^ < 0.1, clumping window > 500 kb). This resulted in 70 DPW SNPs, 61 YOS SNPs, and 39 CPD SNPs (see Figure 1 in main text).

We estimated the total causal effect of CPD on all 11 health outcomes using two-sample, summary level MR (using inverse variance weighted (IVW) regression, MR-Egger, weighted median estimation, and weighted mode estimation). In the MVMR, we estimate the direct causal effect of DPW, YOS, and CPD on all health outcomes using two sample, summary-level MVMR (using MVMR-IVW and MVMR-Egger). We also assessed instrument strength using a conditional F-statistic and instrument validity using Cochran’s Q-statistic.

*Results*

The complete results of the CPD MR and three-exposure MVMR are displayed in Supplementary Tables 3-13. Please note that in the MVMR, the conditional F-statistics indicate that the included SNPs are strongly associated with YOS whilst accounting for DPW and CPD (14.60), but not with DPW whilst accounting for YOS and CPD (5.17), or CPD whilst accounting for DPW and YOS (8.77). The Cochran’s Q statistics were greater than the number of SNPs included in most of the models, indicating heterogeneity.

**STROBE-MR checklist of recommended items to address in reports of Mendelian randomization studies**^1^ ^2^

| **Item No.** | **Section** | **Checklist item** | **Page No.** |
| --- | --- | --- | --- |
| 1 | **TITLE and ABSTRACT** | Indicate Mendelian randomization (MR) as the study’s design in the title and/or the abstract if that is a main purpose of the study | 1, 2 |
|  | **INTRODUCTION** |  |  |
| 2 | **Background** | Explain the scientific background and rationale for the reported study. What is the exposure? Is a potential causal relationship between exposure and outcome plausible? Justify why MR is a helpful method to address the study question | 4,5 |
| 3 | **Objectives** | State specific objectives clearly, including pre-specified causal hypotheses (if any). State that MR is a method that, under specific assumptions, intends to estimate causal effects | 5 |
|  | **METHODS** |  |  |
| 4 | **Study design and data sources** | Present key elements of the study design early in the article. Consider including a table listing sources of data for all phases of the study. For each data source contributing to the analysis, describe the following: |  |
|  | a) | Setting: Describe the study design and the underlying population, if possible. Describe the setting, locations, and relevant dates, including periods of recruitment, exposure, follow-up, and data collection, when available. | 6,7 |
|  | b) | Participants: Give the eligibility criteria, and the sources and methods of selection of participants. Report the sample size, and whether any power or sample size calculations were carried out prior to the main analysis | 6,7 |
|  | c) | Describe measurement, quality control and selection of genetic variants | 6-8 |
|  | d) | For each exposure, outcome, and other relevant variables, describe methods of assessment and diagnostic criteria for diseases | 6-8 |
|  | e) | Provide details of ethics committee approval and participant informed consent, if relevant | - |
| 5 | **Assumptions** | Explicitly state the three core IV assumptions for the main analysis (relevance, independence and exclusion restriction) as well assumptions for any additional or sensitivity analysis | 5 |
| 6 | **Statistical methods: main analysis** | Describe statistical methods and statistics used |  |
|  | a) | Describe how quantitative variables were handled in the analyses (i.e., scale, units, model) | 6-8 |
|  | b) | Describe how genetic variants were handled in the analyses and, if applicable, how their weights were selected | 10,11 |
|  | c) | Describe the MR estimator (e.g. two-stage least squares, Wald ratio) and related statistics. Detail the included covariates and, in case of two-sample MR, whether the same covariate set was used for adjustment in the two samples | 11 |
|  | d) | Explain how missing data were addressed | - |
|  | e) | If applicable, indicate how multiple testing was addressed | - |
| 7 | **Assessment of assumptions** | Describe any methods or prior knowledge used to assess the assumptions or justify their validity | 11 |
| 8 | **Sensitivity analyses and additional analyses** | Describe any sensitivity analyses or additional analyses performed (e.g. comparison of effect estimates from different approaches, independent replication, bias analytic techniques, validation of instruments, simulations) | 11 |
| 9 | **Software and pre-registration** |  |  |
|  | a) | Name statistical software and package(s), including version and settings used | 10 |
|  | b) | State whether the study protocol and details were pre-registered (as well as when and where) | 6 |
|  | **RESULTS** |  |  |
| 10 | **Descriptive data** |  |  |
|  | a) | Report the numbers of individuals at each stage of included studies and reasons for exclusion. Consider use of a flow diagram | 9 (Figure 1) |
|  | b) | Report summary statistics for phenotypic exposure(s), outcome(s), and other relevant variables (e.g. means, SDs, proportions) | - |
|  | c) | If the data sources include meta-analyses of previous studies, provide the assessments of heterogeneity across these studies | - |
|  | d) | For two-sample MR:  i.  Provide justification of the similarity of the genetic variant-exposure associations between the exposure and outcome samples  ii.  Provide information on the number of individuals who overlap between the exposure and outcome studies | 7 |
| 11 | **Main results** |  |  |
|  | a) | Report the associations between genetic variant and exposure, and between genetic variant and outcome, preferably on an interpretable scale | 11-16 |
|  | b) | Report MR estimates of the relationship between exposure and outcome, and the measures of uncertainty from the MR analysis, on an interpretable scale, such as odds ratio or relative risk per SD difference | 11-16 |
|  | c) | If relevant, consider translating estimates of relative risk into absolute risk for a meaningful time period | - |
|  | d) | Consider plots to visualize results (e.g. forest plot, scatterplot of associations between genetic variants and outcome versus between genetic variants and exposure) | 13 (Figure 2) |
| 12 | **Assessment of assumptions** |  |  |
|  | a) | Report the assessment of the validity of the assumptions | 11-16 |
|  | b) | Report any additional statistics (e.g., assessments of heterogeneity across genetic variants, such as *I^2^*, Q statistic or E-value) | 12 |
| 13 | **Sensitivity analyses and additional analyses** |  |  |
|  | a) | Report any sensitivity analyses to assess the robustness of the main results to violations of the assumptions | 11 |
|  | b) | Report results from other sensitivity analyses or additional analyses | 11 |
|  | c) | Report any assessment of direction of causal relationship (e.g., bidirectional MR) | - |
|  | d) | When relevant, report and compare with estimates from non-MR analyses | - |
|  | e) | Consider additional plots to visualize results (e.g., leave-one-out analyses) | - |
|  | **DISCUSSION** |  |  |
| 14 | **Key results** | Summarize key results with reference to study objectives | 17 |
| 15 | **Limitations** | Discuss limitations of the study, taking into account the validity of the IV assumptions, other sources of potential bias, and imprecision. Discuss both direction and magnitude of any potential bias and any efforts to address them | 19,20 |
| 16 | **Interpretation** |  |  |
|  | a) | Meaning: Give a cautious overall interpretation of results in the context of their limitations and in comparison with other studies | 20,21 |
|  | b) | Mechanism: Discuss underlying biological mechanisms that could drive a potential causal relationship between the investigated exposure and the outcome, and whether the gene-environment equivalence assumption is reasonable. Use causal language carefully, clarifying that IV estimates may provide causal effects only under certain assumptions | 20,21 |
|  | c) | Clinical relevance: Discuss whether the results have clinical or public policy relevance, and to what extent they inform effect sizes of possible interventions | - |
| 17 | **Generalizability** | Discuss the generalizability of the study results (a) to other populations, (b) across other exposure periods/timings, and (c) across other levels of exposure | 20,21 |
|  | **OTHER INFORMATION** |  |  |
| 18 | **Funding** | Describe sources of funding and the role of funders in the present study and, if applicable, sources of funding for the databases and original study or studies on which the present study is based | 22 |
| 19 | **Data and data sharing** | Provide the data used to perform all analyses or report where and how the data can be accessed, and reference these sources in the article. Provide the statistical code needed to reproduce the results in the article, or report whether the code is publicly accessible and if so, where | 6-8 |
| 20 | **Conflicts of Interest** | All authors should declare all potential conflicts of interest | 22 |

This checklist is copyrighted by the Equator Network under the Creative Commons Attribution 3.0 Unported (CC BY 3.0) license.

1. Skrivankova VW, Richmond RC, Woolf BAR, Yarmolinsky J, Davies NM, Swanson SA, et al. Strengthening the Reporting of Observational Studies in Epidemiology using Mendelian Randomization (STROBE-MR) Statement. JAMA. 2021;under review.
2. Skrivankova VW, Richmond RC, Woolf BAR, Davies NM, Swanson SA, VanderWeele TJ, et al. Strengthening the Reporting of Observational Studies in Epidemiology using Mendelian Randomisation (STROBE-MR): Explanation and Elaboration. BMJ. 2021;375:n2233.
